# Online speech synthesis using a chronically implanted brain-computer interface in an individual with ALS

**DOI:** 10.1101/2023.06.30.23291352

**Authors:** Miguel Angrick, Shiyu Luo, Qinwan Rabbani, Daniel N. Candrea, Samyak Shah, Griffin W. Milsap, William S. Anderson, Chad R. Gordon, Kathryn R. Rosenblatt, Lora Clawson, Nicholas Maragakis, Francesco V. Tenore, Matthew S. Fifer, Hynek Hermansky, Nick F. Ramsey, Nathan E. Crone

## Abstract

Recent studies have shown that speech can be reconstructed and synthesized using only brain activity recorded with intracranial electrodes, but until now this has only been done using retrospective analyses of recordings from able-bodied patients temporarily implanted with electrodes for epilepsy surgery. Here, we report online synthesis of intelligible words using a chronically implanted brain-computer interface (BCI) in a clinical trial participant (ClinicalTrials.gov, NCT03567213) with dysarthria due to amyotrophic lateral sclerosis (ALS). We demonstrate a reliable BCI that synthesizes commands freely chosen and spoken by the user from a vocabulary of 6 keywords originally designed to allow intuitive selection of items on a communication board. Our results show for the first time that a speech-impaired individual with ALS can use a chronically implanted BCI to reliably produce synthesized words that are intelligible to human listeners while preserving the participants voice profile.

## Introduction

A variety of neurological disorders, including amyotrophic lateral sclerosis (ALS), can severely affect speech production and other purposeful movements while sparing cognition. This can result in varying degrees of communication impairments, including Locked-In Syndrome (LIS) (Bauer, 1997; Smith, 2005), in which patients can only answer yes/no questions or select from sequentially presented options using eyeblinks, eye movements, or other residual movements. Individuals such as these may use augmentative and alternative technologies (AAT) to select among options on a communication board, but this communication can be slow, effortful, and may require caregiver intervention. Recent advances in implantable brain-computer interfaces (BCIs) have demonstrated the feasibility of establishing and maintaining communication using a variety of direct brain control strategies that bypass weak muscles, for example to control a switch scanner (Vansteensel, 2016; Chaudhary, 2022), a computer cursor (Pandarinath, 2017), to write letters (Willet, 2021) or to spell words using a hybrid approach of eye-tracking and attempted movement detection (Oxley, 2021). However, these communication modalities are still slower, more effortful, and less intuitive than speech-based BCI control (Chang, 2020).

Recent studies have also explored the feasibility of decoding attempted speech from brain activity, outputting text or even acoustic speech, which could potentially carry more linguistic information such as intonation and prosody. Previous studies have reconstructed acoustic speech in offline analysis from linear regression models (Herff, 2016), convolutional (Angrick, 2019) and recurrent neural networks (Anumanchipalli, 2019; Wairagkar, 2023), and encoder-decoder architectures (Kohler, 2022). Concatenative approaches from the text-to-speech synthesis domain have also been explored (Herff, 2019; Wilson, 2020), and voice activity has been identified in electrocorticographic (ECoG) (Kanas, 2014) and stereotactic EEG recordings (Soroush, 2021). Moreover, speech decoding has been performed at the level of American English phonemes (Mugler, 2014), spoken vowels (Bouchard, 2013; Bouchard, 2014), spoken words (Kellis, 2010) and articulatory gestures (Mugler, 2015; Mugler, 2018).

Until now, brain-to-speech decoding has primarily been reported in individuals with unimpaired speech, such as patients temporarily implanted with intracranial electrodes for epilepsy surgery. To date, it is unclear to what extent these findings will ultimately translate to individuals with motor speech impairments, as in ALS and other neurological disorders. Recent studies have demonstrated how neural activity acquired from an ECoG grid (Moses, 2021) or from microelectrodes (Willett, 2023) can be used to recover text from a patient with anarthria due to a brainstem stroke, or from a patient with dysarthria due to ALS, respectively. Prior to these studies, a landmark study allowed a locked-in volunteer to control a real-time synthesizer generating vowel sounds (Guenther, 2009). However, there have been no reports to date of direct closed-loop synthesis of intelligible spoken words.

Here, we show that an individual with ALS participating in a clinical trial of an implantable BCI (ClinicalTrials.gov, NCT03567213) was able to produce audible, intelligible words that closely resembled his own voice, spoken at his own pace. Speech synthesis was accomplished through online decoding of ECoG signals generated during overt speech production from cortical regions previously shown to represent articulation and phonation, following similar previous work (Bouchard, 2013; Chartier, 2018; Anumanchipalli, 2019; Akbari, 2019). Our participant had considerable impairments in articulation and phonation. He was still able to produce some words that were intelligible when spoken in isolation, but his sentences were often unintelligible. Here, we focused on a closed vocabulary of 6 keywords, chosen for intuitive navigation of a communication board. Our participant was capable of producing these 6 keywords individually with a high degree of intelligibility. We acquired training data over a period of 6 weeks and deployed the speech synthesis BCI in several separate closed-loop sessions. Since the participant could still produce speech, we were able to time-align the individual’s neural and acoustic signals to enable a mapping between his cortical activity during overt speech production processes and his voice’s acoustic features. We chose to provide delayed rather than simultaneous auditory feedback in anticipation of ongoing deterioration in the patient’s speech due to ALS, with increasing discordance and interference between actual and BCI-synthesized speech. This design choice would be ideal for a neuroprosthetic device that remains capable of producing intelligible words as an individual’s speech becomes increasingly unintelligible.

Here, we present the first closed-loop, self-paced BCI that translates brain activity to acoustic speech that resembles characteristics of the user’s voice profile, with most synthesized words of sufficient intelligibility to be correctly recognized by human listeners. This work makes an important step in translating recent results from speech synthesis from neural signals to their intended users with neurological speech impairments, by first focusing on a closed vocabulary that the participant can reliably generate at his own pace.

### Approach

In order to synthesize acoustic speech from neural signals, we designed a pipeline that consisted of three recurrent neural networks (RNNs) to 1) identify and buffer speech-related neural activity, 2) transform sequences of speech-related neural activity into an intermediate acoustic representation, and 3) eventually recover the acoustic waveform using a vocoder. Figure 1 shows a schematic overview of our approach. We acquired ECoG signals from two electrode grids that covered cortical representations for speech production including ventral sensorimotor cortex and the dorsal laryngeal area (Figure 1.A). Here, we focused only on a subset of electrodes that had previously been identified as showing significant changes in high-gamma activity associated with overt speech production (see Supplementary Figure 2). From the raw ECoG signals, our closed-loop speech synthesizer extracted broadband high-gamma power features (70-170 Hz) that had previously been demonstrated to encode speech-related information useful for decoding speech (Figure 1.B) (Herff, 2019; Angrick, 2019).

**Figure 1.**
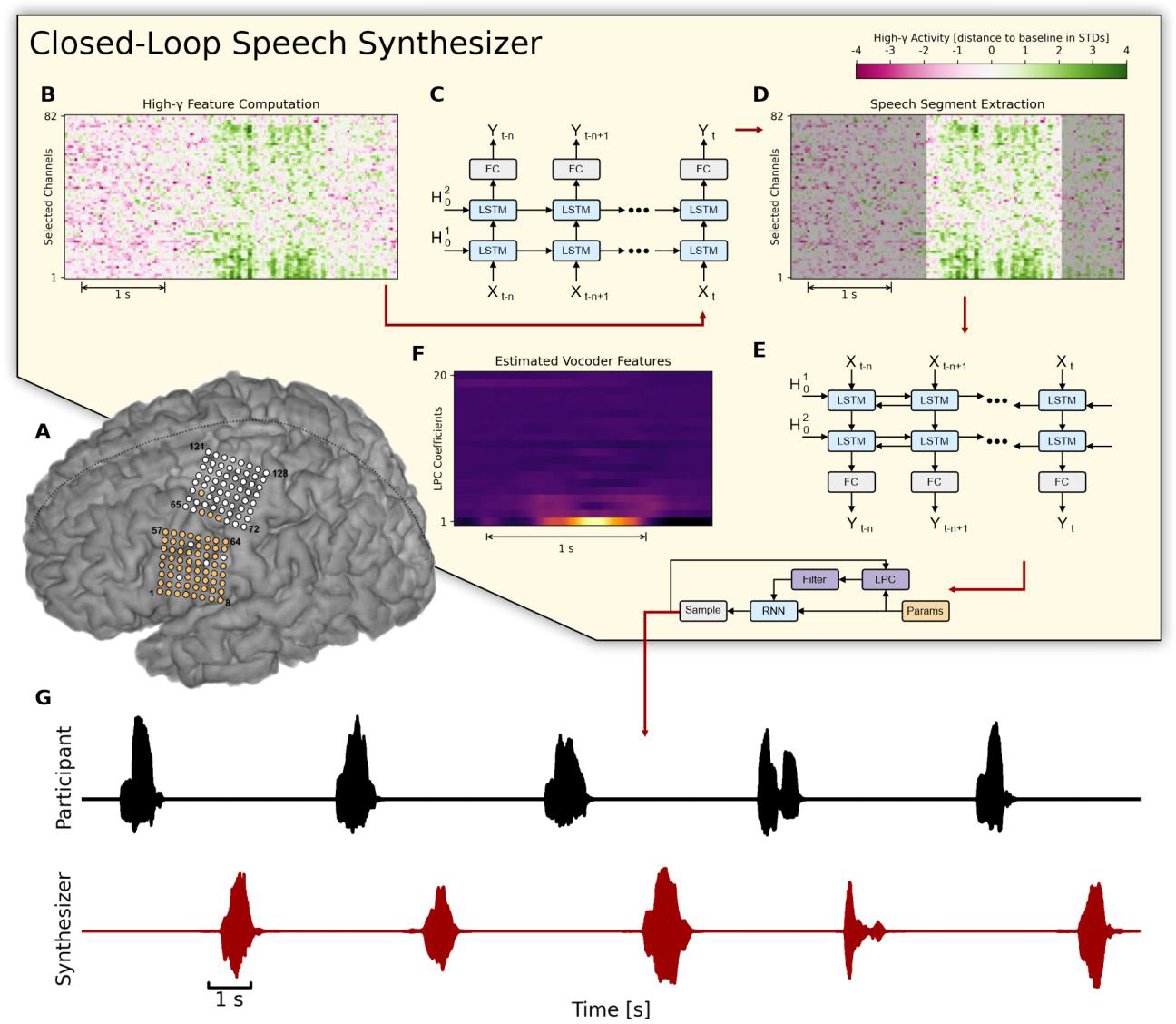
| Overview of the closed-loop speech synthesizer. (**A**) Neural activity is acquired from a subset of 64 electrodes (highlighted in orange) from two 8 x 8 ECoG electrode arrays covering sensorimotor areas for face and tongue, and for upper limb regions. (**B**) The closed-loop speech synthesizer extracts high-gamma features to reveal speech-related neural correlates of attempted speech production and propagates each frame to a neural voice activity detection (nVAD) model (**C**) that identifies and extracts speech segments (**D**). When the participant finishes speaking a word, the nVAD model forwards the high-gamma activity of the whole extracted sequence to a bidirectional decoding model (**E**) which estimates acoustic features (**F**) that can be transformed into an acoustic speech signal. (**G**) The synthesized speech is played back as acoustic feedback.

We used a unidirectional RNN to identify and buffer sequences of high-gamma activity frames and extract speech segments (Figure 1.C-D). This neural voice activity detection (nVAD) model internally employed a strategy to correct misclassified frames based on each frame’s temporal context, and additionally included a context window of 0.5 s to allow for smoother transitions between speech and non-speech frames. Each buffered sequence was forwarded to a bidirectional decoding model that mapped high-gamma features onto 18 Bark-scale cepstral coefficients (Moore, 2012) and 2 pitch parameters, henceforth referred to as LPC coefficients (Taylor, 2009; Valin, 2019) (Figure 1.E-F). We used a bidirectional architecture to include past and future information while making frame-wise predictions. Estimated LPC coefficients were transformed into an acoustic speech signal using the LPCNet vocoder (Valin, 2019) and played back as delayed auditory feedback (Figure 1.G).

## Results

### Synthesis Performance

When deployed in sessions with the participant for online decoding, our speech-synthesis BCI was reliably capable of producing acoustic speech that captured many details and characteristics of the voice and pacing of the participant’s natural speech, often yielding a close resemblance to the words spoken in isolation from the participant. Figure 2.A provides examples of original and synthesized waveforms for a representative selection of words time-aligned by subtracting the duration of the extracted speech segment from the nVAD. Onset timings from the reconstructed waveforms indicate that the decoding model captured the flow of the spoken word while also synthesizing silence around utterances for smoother transitions. Figure 2.B shows the corresponding acoustic spectrograms for the spoken and synthesized words, respectively. The spectral structures of the original and synthesized speech shared many common characteristics and achieved average correlation scores of 0.67 (±0.18 standard deviation) suggesting that phoneme and formant-specific information were preserved.

**Figure 2.**
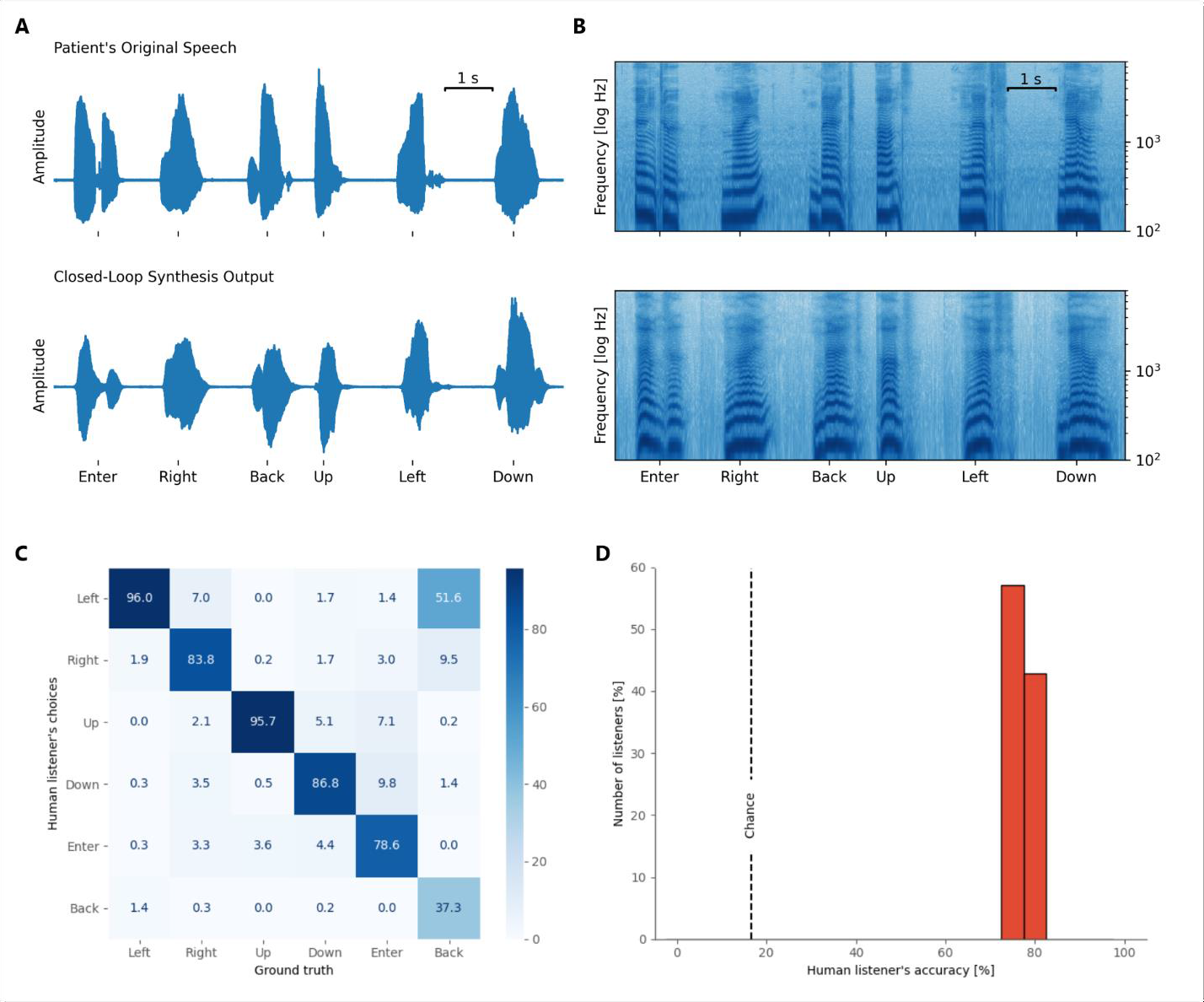
| Evaluation of the synthesized words. (**A**) Visual example of time-aligned original and reconstructed acoustic speech waveforms and their spectral representations (**B**) for 6 words that were recorded during one of the closed-loop sessions. Speech spectrograms are shown between 100 and 8000 Hz with a logarithmic frequency range to emphasize formant frequencies. (**C**) The confusion matrix between human listeners and ground truth. (**D**) Distribution of accuracy scores from all who performed the listening test for the synthesized speech samples. Dashed line shows chance performance (16.7%).

We conducted 3 sessions across 3 different days (approximately 5 and a half months after the training data was acquired, each session lasted 6 min) to repeat the experiment with acoustic feedback from the BCI to the participant (see Supplementary Video 1 for an excerpt). Other experiment parameters were not changed. All synthesized words were played back on loudspeakers while simultaneously recorded for evaluation.

To assess the intelligibility of the synthesized words, we conducted listening tests in which human listeners played back individual samples of the synthesized words and selected the word that most closely resembled each sample. Additionally, we mixed in samples that contained the originally spoken words. This allowed us to assess the quality of the participant’s natural speech. We recruited a cohort of 21 native English speakers to listen to all samples that were produced during our 3 closed-loop sessions. Out of 180 samples, we excluded 2 words because the nVAD model did not detect speech activity and therefore no speech output was produced by the decoding model. We also excluded a few cases where speech activity was falsely detected by the nVAD model, which resulted in synthesized silence and remained unnoticed to the participant.

Overall, human listeners achieved an accuracy score of 80%, indicating that the majority of synthesized words could be correctly and reliably recognized. Figure 2.C presents the confusion matrix regarding only the synthesized samples where the ground truth labels and human listener choices are displayed on the X- and Y-axes respectively. The confusion matrix shows that human listeners were able to recognize all but one word at very high rates. “Back” was recognized at low rates, albeit still above chance, and was most often mistaken for “Left”. This could have been due in part to the close proximity of the vowel formant frequencies for these two words. The participant’s weak tongue movements may have deemphasized the acoustic discriminability of these words, in turn resulting in the vocoder synthesizing a version of “back” that was often indistinct from “left”. In contrast, the confusion matrix also shows that human listeners were confident in distinguishing the words “Up” and “Left”. The decoder synthesized an intelligible but incorrect word in only 4% of the cases, and all listeners accurately recognized the incorrect word. Note that all keywords in the vocabulary were chosen for intuitive command and control of a computer interface, for example a communication board, and were not designed to be easily discriminable for BCI applications.

Figure 2.D summarizes individual accuracy scores from all human listeners from the listening test in a histogram. All listeners recognized between 75% and 84% of the synthesized words. All human listeners achieved accuracy scores above chance (16.7%). In contrast, when tested on the participant’s natural speech, our human listeners correctly recognized almost all samples of the 6 keywords (99.8%).

### Anatomical and temporal contributions

In order to understand which cortical areas contributed to identification of speech segments, we conducted a saliency analysis (Montavon, 2018) to reveal the underlying dynamics in high-gamma activity changes that explain the binary decisions made by our nVAD model. We utilized a method from the image processing domain (Simonyan, 2014) that queries spatial information indicating which pixels have contributed to a classification task. In our case, this method ranked individual high-gamma features over time by their influence on the predicted speech onsets (PSO). The absolute values of their gradients allowed interpretations of which contributions had the highest or lowest impact on the class scores from anatomical and temporal perspectives.

The general idea is illustrated in Figure 3.B. In a forward pass, we first estimated for each trial the PSO by propagating through each time step until the nVAD model made a positive prediction. From here, we then applied backpropagation through time to compute all gradients with respect to the model’s input high-gamma features. Relevance scores |R| were computed by taking the absolute value of each partial derivative and the maximum value across time was used as the final score for each electrode (Simonyan, 2014). Note that we only performed backpropagation through time for each PSO, and not for whole speech segments.

**Figure 3.**
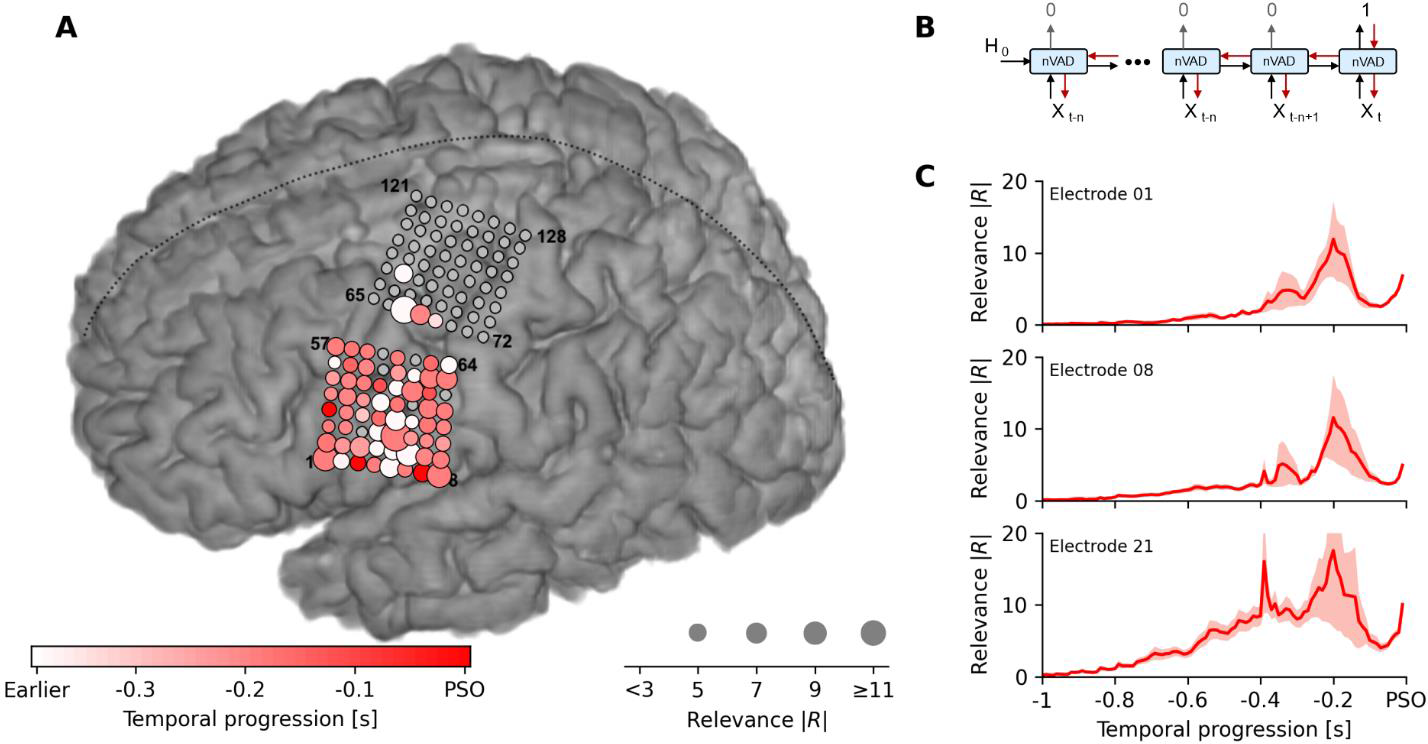
| Changes in high-gamma activity across motor, premotor and somatosensory cortices trigger detection of speech output. **(A)** Saliency analysis shows that changes in high-gamma activity predominantly from 300 to 100 ms prior to predicted speech onset (PSO) strongly influenced the nVAD model’s decision. Electrodes covering motor, premotor and somatosensory cortices show the impact of model decisions, while electrodes covering the dorsal laryngeal area only modestly added information to the prediction. Grey electrodes were either not used, bad channels or had no notable contributions. **(B)** Illustration of the general procedure on how relevance scores were computed. For each time step *t*, relevance scores were computed by backpropagation through time across all previous high-gamma frames *X_t_*. Predictions of 0 correspond to no-speech, while 1 represents speech frames. **(C)** Temporal progression of mean magnitudes of the absolute relevance score in 3 selected channels that strongly contributed to PSOs. Shaded areas reflect the standard error of the mean (N=60). Units of the relevance scores are in 10^−3^.

Results from the saliency analysis are shown in Figure 3.A. For each channel, we display the PSO-specific relevance scores by encoding the maximum magnitude of the influence in the size of the circles (bigger circles mean stronger influence on the predictions), and the temporal occurrence of that maximum in the respective color coding (lighter electrodes have their maximal influence on the PSO earlier). The color bar at the bottom limits the temporal influence to −400 ms prior to PSO, consistent with previous reports about speech planning (Indefrey, 2011) and articulatory representations (Bouchard, 2013). The saliency analysis showed that the nVAD model relied on a broad network of electrodes covering motor, premotor and somatosensory cortices whose collective changes in the high-gamma activity were relevant for identifying speech. Meanwhile, voice activity information encoded in the dorsal laryngeal area (highlighted electrodes in the upper grid in Figure 3.A) (Bouchard, 2013) only mildly contributed to the PSO.

Figure 3.C shows relevance scores over a time period of 1 s prior to PSO for 3 selected electrodes that strongly contributed to predicting speech onsets. In conjunction with the color coding from Figure 3.A, the temporal associations were consistent with previous studies that examined phoneme decoding over fixed window sizes of 400 ms (Mugler, 2014) and 500 ms (Ramsey, 2018; Jiang, 2016) around speech onset times, suggesting that the nVAD model benefited from neural activity during speech planning and phonological processing (Indefrey, 2011) when identifying speech onset. We hypothesize that the decline in the relevance scores after −200 ms can be explained by the fact that voice activity information might have already been stored in the long short-term memory of the nVAD model and thus changes in neural activity beyond this time had less influence on the prediction.

## Discussion

Here we demonstrate the feasibility of a closed-loop BCI that is capable of online synthesis of intelligible words using intracranial recordings from the speech cortex of an ALS participant. Recent studies (Anumanchipalli, 2019; Angrick, 2019; Kohler, 2022) suggest that deep learning techniques are a viable tool to reconstruct acoustic speech from ECoG signals. We found an approach consisting of three consecutive RNN architectures that identify and transform neural speech correlates into an acoustic waveform that can be streamed over the loudspeaker as neurofeedback, resulting in an 80% intelligibility score on a closed-vocabulary, keyword reading task.

The majority of human listeners were able to correctly recognize most synthesized words – a significant advance to the field given that intelligible speech synthesis has so far been limited to offline analyses. All words from the closed vocabulary were chosen based on intuitive real-life applicability rather than being constructed to elicit discriminable neural activity that benefits decoder performance. The listening tests suggest that the words “Left” and “Back” were responsible for the majority of misclassified words. These words share very similar articulatory features, and our participant’s speech impairments likely made these words less discriminable in the synthesis process.

Saliency analysis showed that our nVAD approach used information encoded in the high-gamma band across predominantly motor, premotor and somatosensory cortices, while electrodes covering the dorsal laryngeal area only marginally contributed to the identification of speech onsets. In particular, neural changes previously reported to be important for speech planning and phonological processing (Indefrey, 2011; Bouchard, 2013) appeared to have a profound impact. Here, the analysis indicates that our nVAD model learned a proper representation of spoken speech processes, providing a connection between neural patterns learned by the model and the spatio-temporal dynamics of speech production.

Our participant was chronically implanted with 128 subdural ECoG electrodes, roughly half of which covered cortical areas where similar high-gamma responses have been reliably elicited during overt speech (Crone, 2001; Mugler, 2014; Ramsey, 2018; Bouchard, 2013) and have been used for offline decoding and reconstruction of speech (Angrick, 2019; Anumanchipalli, 2019). This study and others like it (Moses, 2021; Moses, 2019; Herff, 2015) explored the potential of ECoG-based BCIs to augment communication for individuals with motor speech impairments due to a variety of neurological disorders, including ALS and brainstem stroke. A potential advantage of ECoG for BCI is the stability of signal quality over long periods of time (Morrell, 2011). In a previous study of an individual with locked-in syndrome due to ALS, a fully implantable ECoG BCI with fewer electrodes provided a stable switch for a spelling application over a period of more than 3 years (Pels, 2019). Similarly, Rao et al. reported robust responses for ECoG recordings over the speech-auditory cortex for two drug-resistant epilepsy patients over a period of 1.5 years (Rao, 2017). The speech synthesis approach we demonstrated here used training data from 5 and a half months prior to testing and produced similar results over 3 separate days of testing, with recalibration but no retraining in each session. These findings suggest that the correspondence between neural activity in ventral sensorimotor cortex and speech acoustics were not significantly changed over this time period. Although longitudinal testing over longer time periods will be needed to explicitly test this, our findings provide additional support for the stability of ECoG as a BCI signal source for speech synthesis.

Our approach used a speech synthesis model trained on neural data acquired during overt speech production. This constrains our current approach to patients with speech motor impairments in which vocalization is still possible and in which speech may still be intelligible. Given the increasing use of voice banking among people living with ALS, it may also be possible to improve the intelligibility of synthetic speech using an approach similar to ours, even in participants with unintelligible or absent speech. This speech could be utilized as a surrogate but would require careful alignment to speech attempts. Likewise, the same approach could be used with a generic voice, though this would not preserve the individual’s speech characteristics. Nevertheless, it remains to be seen how long our approach will continue to produce intelligible speech as our patient’s neural responses and articulatory impairments change over time due to ALS. Previous studies of long-term ECoG signal stability and BCI performance in patients with more severe motor impairments suggest that this may be possible (Vansteensel, 2016; Silversmith, 2021).

Although our approach allowed for online, closed-loop production of synthetic speech that preserved our participant’s individual voice characteristics, the bidirectional LSTM imposed a delay in the audible feedback until after the patient spoke each word. We considered this delay to be not only acceptable, but potentially desirable, given our patient’s speech impairments and the likelihood of these impairments worsening in the future due to ALS. Although normal speakers use immediate acoustic feedback to tune their speech motor output (Denes, 1993), individuals with progressive motor speech impairments are likely to reach a point at which there is a significant, and distracting, mismatch between the subject’s speech and the synthetic speech produced by the BCI. In contrast, providing acoustic feedback immediately after each utterance gives the user clear and uninterrupted output that they can use to improve subsequent speech attempts, if necessary.

While our results are promising, the approach used here did not allow for synthesis of unseen words. The bidirectional architecture of the decoding model learned variations of the neural dynamics of each word and was capable of recovering their acoustic representations from corresponding sequences of high-gamma frames. This approach does not capture more fine-grained and isolated part-of-speech units, such as syllables or phonemes. However, previous research (Anumanchipalli, 2019) has shown that speech synthesis approaches based on bidirectional architectures can generalize to unseen elements that were not part of the training set. Future research will be needed to expand the currently limited vocabulary for speech synthesis, and to explore to what extent similar or different approaches are able to extrapolate to words that are not in the vocabulary of the training set.

Our demonstration here builds on previous seminal studies of the cortical representations for articulation and phonation (Chartier, 2018; Ramsey, 2018; Bouchard, 2013) in epilepsy patients implanted with similar subdural ECoG arrays for less than 30 days. These studies and others using intraoperative recordings have also supported the feasibility of producing synthetic speech from ECoG high-gamma responses (Anumanchipalli, 2019; Angrick, 2019; Akbari, 2019), but these demonstrations were based on offline analysis of ECoG signals that were previously recorded in subjects with normal speech. Here, a participant with impaired articulation and phonation was able to use a chronically implanted investigational device to produce acoustic speech that retained his unique voice characteristics. This was made possible through online decoding of ECoG high-gamma responses, using an algorithm trained on data collected months before. Notwithstanding the current limitations of our approach, our findings here provide a promising proof-of-concept that ECoG BCIs utilizing online speech synthesis can serve as alternative and augmentative communication devices for people living with ALS, as well as others with speech impairments due to paralysis. Moreover, our findings should motivate continued research on the feasibility of using BCIs to preserve or restore vocal communication in clinical populations where this is needed.

## Materials and Methods

### Participant

Our participant was a male native English speaker in his 60s with ALS who was enrolled in a clinical trial (NCT03567213), approved by the Johns Hopkins University Institutional Review Board (IRB) and by the FDA (under an investigational device exemption) to test the safety and preliminary efficacy of a brain-computer interface composed of subdural electrodes and a percutaneous connection to external EEG amplifiers and computers. Diagnosed with ALS 8 years prior to implantation, our participant’s motor impairments had chiefly affected bulbar and upper extremity muscles and had resulted in motor impairments sufficient to render continuous speech mostly unintelligible (though individual words were intelligible), and to require assistance with most activities of daily living. The participant gave consent after being informed of the nature of the research and implant-related risks and was implanted with the study device in July 2022.

### Study device and implantation

The study device was composed of two 8 x 8 subdural electrode grids (PMT Corporation, Chanhassen, MN) connected to a percutaneous 128-channel Neuroport pedestal (Blackrock Neurotech, Salt Lake City, UT). Both subdural grids contained platinum-iridium disc electrodes (0.76 mm thickness, 2-mm diameter exposed surface) with 4 mm center-to-center spacing and a total surface area of 12.11 cm^2^ (36.6 mm x 33.1 mm).

The study device was surgically implanted during a craniotomy with the ECoG grids placed on the pial surface of sensorimotor representations for speech and upper extremity movements in the left hemisphere. Careful attention was made to assure that the scalp flap incision was well away from the external pedestal. Cortical representations were targeted using anatomical landmarks from pre-operative structural (MRI) and functional imaging (fMRI), in addition to somatosensory evoked potentials measured intraoperatively. Two reference wires attached to the Neuroport pedestal were implanted in the subdural space on the outward facing surface of the subdural grids. The participant was awoken during the craniotomy to confirm proper functioning of the study device and final placement of the two subdural grids. For this purpose, the participant was asked to repeatedly speak a single word as event-related ECoG spectral responses were noted. On the same day, the participant had a post-operative CT which was then co-registered to a pre-operative MRI to verify the anatomical locations of the two grids.

### Data Recording

During all training and testing sessions, the Neuroport pedestal was connected to a 128-channel NeuroPlex-E headstage that was in turn connected by a mini-HDMI cable to a NeuroPort Biopotential Signal Processor (Blackrock Neurotech, Salt Lake City, UT, USA) and external computers.

Acoustic speech was recorded through an external microphone (BETA® 58A, SHURE, Niles, IL) in a room isolated from external acoustic and electronic noise, then amplified and digitized by an external audio interface (H6-audio-recorder, Zoom Corporation, Tokyo, Japan). The acoustic speech signal was split and forwarded to: 1) an analog input of the NeuroPort Biopotential Signal Processor (NSP) to be recorded at the same frequency and in synchrony with the neural signals, and 2) the testing computer to capture high-quality (48 kHz) recordings. We applied cross-correlation to align the high-quality recordings with the synchronized audio signal from the NSP.

### Experiment Recordings & Task design

Each recording day began with a syllable repetition task to acquire cortical activity to be used for baseline normalization. Each syllable was audibly presented through a loudspeaker, and the participant was instructed to recite the heard stimulus by repeating it aloud. Stimulus presentation lasted for 1 s, and trial duration was set randomly in the range of 2.5 s and 3.5 s with a step size of 80 ms. In the syllable repetition task, the participant was instructed to repeat 12 consonant-vowel syllables (Supplementary Table 1), in which each syllable was repeated 5 times. We extracted high-gamma frames from all trials to compute for each day the mean and standard deviation statistics for channel-specific normalization.

To collect data for training our nVAD and speech decoding model, we recorded ECoG during multiple blocks of a speech production task over a period of 6 weeks. During the task, the participant read aloud single words that were prompted on a computer screen, interrupted occasionally by a silence trial in which the participant was instructed to say nothing. The words came from a closed vocabulary of 6 words ("Left", "Right", "Up", "Down", "Enter", "Back", and “…” for silence). In each block, there were five repetitions of each word (60 words in total) that appeared in a pseudo-randomized order by having a fixed set of seeds to control randomization orders. Each word was shown for 2 s per trial with an intertrial interval of 3 s. The participant was instructed to read the prompted word aloud as soon as it appeared. Because his speech was slow, effortful, and dysarthric, the participant may have sometimes used some of the intertrial interval to complete word production. However, offline analysis verified at least 1 s between the end of each spoken word and the beginning of the next trial, assuring that enough time had passed to avoid ECoG high-gamma responses leaking into subsequent trials. In each block, neural signals and audibly vocalized speech were acquired in parallel and stored to disc using BCI2000 (Schalk, 2004).

We recorded training, validation, and test data for 10 days, and deployed our approach for synthesizing speech online 5 and a half months later. During the online task, the synthesized output was played to the participant while he performed the same keyword reading task as in the training sessions. The feedback from each synthesized word began after he spoke the same word, avoiding any interference with production from the acoustic feedback. The validation dataset was used for finding appropriate hyperparameters to train both nVAD and the decoding model. The test set was used to validate final model generalizability before online sessions. We also used the test set for the saliency analysis. In total, the training set was comprised of 1,570 trials that aggregated to approximately 80 min of data (21.8 min are pure speech), while the validation and test set contained 70 trials each with around 3 min of data (0.9 min pure speech), respectively. The data in each of these datasets were collected on different days, so that no baseline or other statistics in the training set leaked into the validation or test set.

### Signal Processing & Feature Extraction

Neural signals were transformed into broadband high-gamma power features that have been previously reported to closely track the timing and location of cortical activation during speech and language processes (Crone, 2001; Leuthardt, 2012). In this feature extraction process, we first re-referenced all channels within each 64-contact grid to a common-average reference (CAR filtering), excluding channels with poor signal quality in any training session. Next, we selected all channels that had previously shown significant high-gamma responses during the syllable repetition task described above. This included 64 channels (Supplementary Figure 2, channels with blue outlines) across motor, premotor and somatosensory cortices, including the dorsal laryngeal area. From here, we applied two IIR Butterworth filters (both with filter order 8) to extract the high-gamma band in the range of 70 to 170 Hz while subsequently attenuating the first harmonic (118-122 Hz) of the line noise. For each channel, we computed logarithmic power features based on windows with a fixed length of 50 ms and a frameshift of 10 ms. To estimate speech-related increases in broadband high-gamma power, we normalized each feature by the day-specific statistics of the high-gamma power features accumulated from the syllable repetition task.

For the acoustic recordings of the participant’s speech, we downsampled the time-aligned high-quality microphone recordings from 48 kHz to 16 kHz. From here, we padded the acoustic data by 16 ms to account for the shift introduced by the two filters on the neural data and estimated the boundaries of speech segments using an energy-based voice activity detection algorithm (Povey, 2011). Likewise, we computed acoustic features in the LPC coefficient space through the encoding functionality of the LPCNet vocoder. Both voice activity detection and LPC feature encoding were configured to operate on 10 ms frameshifts to match the number of samples from the broadband high-gamma feature extraction pipeline.

### Network Architectures

Our proposed approach relied on three recurrent neural network architectures: 1) a unidirectional model that identified speech segments from the neural data, 2) a bidirectional model that translated sequences of speech-related high-gamma activity into corresponding sequences of LPC coefficients representing acoustic information, and 3) LPCNet (Valin, 2019), which converted those LPC coefficients into an acoustic speech signal.

The network architecture of the unidirectional nVAD model was inspired by Zen et al. (Zen, 2015) in using a stack of two LSTM layers with 150 units each, followed by a linear fully connected output layer with two units representing speech or non-speech class target logits (Figure 4). We trained the unidirectional nVAD model using truncated backpropagation through time (BPTT) (Sutskever, 2013) to keep the costs of single parameter updates manageable. We initialized this algorithm’s hyperparameters *k_1_* and *k_2_* to 50 and 100 frames of high-gamma activity, respectively, such that the unfolding procedure of the backpropagation step was limited to 100 frames (1 s) and repeated every 50 frames (500 ms). Dropout was used as a regularization method with a probability of 50% to counter overfitting effects (Srivastava, 2014). Comparison between predicted and target labels was determined by the cross-entropy loss. We limited the network training using an early stopping mechanism that evaluated after each epoch the network performance on a held-out validation set and kept track of the best model weights by storing the model weights only when the frame-wise accuracy score was bigger than before. The learning rate of the stochastic gradient descent optimizer was dynamically adjusted in accordance with the RMSprop formula (Ruder, 2016) with an initial learning rate of 0.001. Using this procedure, the unidirectional nVAD model was trained for 27,975 update steps, achieving a frame-wise accuracy of 93.4% on held-out validation data. The architecture of the nVAD model had 311,102 trainable weights.

**Figure 4.**
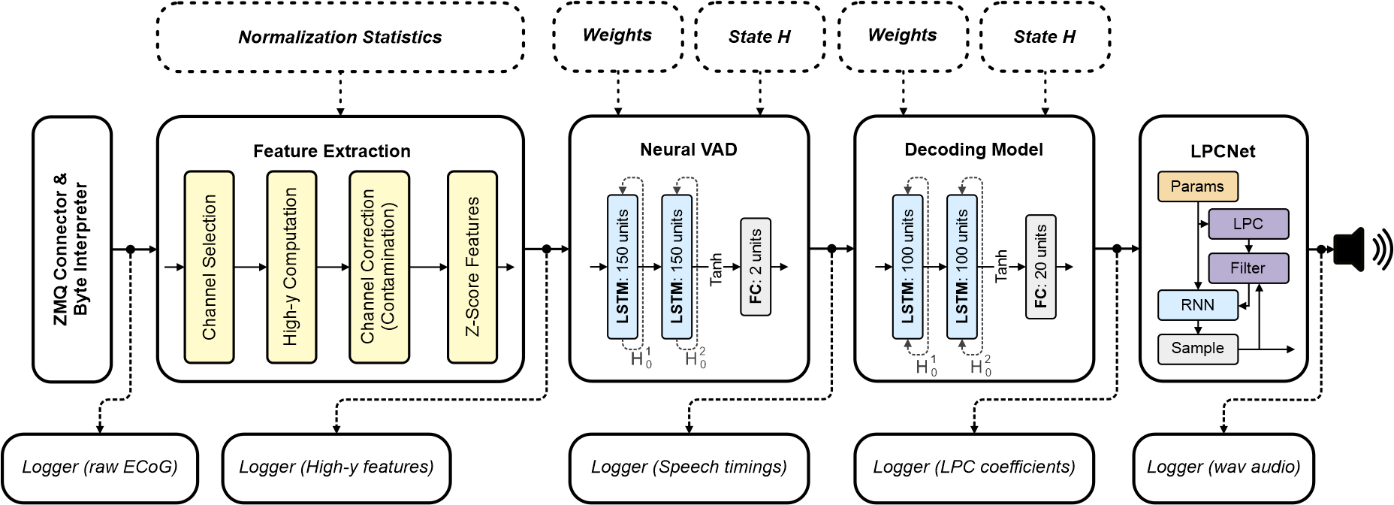
| System overview of the closed-loop architecture. The computational graph is designed as a directed acyclic network. Solid shapes represent ezmsg units, dotted ones represent initialization parameters. Each unit is responsible for a self-contained task and distributes their output to all its subscribers. Logger units run in separate processes to not interrupt the main processing chain for synthesizing speech.

The network architecture of the bidirectional decoding model had a very similar configuration to the unidirectional nVAD but employed a stack of bidirectional LSTM layers for sequence modelling (Anumanchipalli, 2019) to include past and future contexts. Since the acoustic space of the LPC components was continuous, we used a linear fully connected output layer for this regression task. Figure 4 contains an illustration of the network architecture of the decoding model. In contrast to the unidirectional nVAD model, we use standard BPTT to account for both past and future contexts within each extracted segment identified as spoken speech. The architecture of the decoding model had 378,420 trainable weights and was trained for 14,130 update steps using a stochastic gradient descent optimizer. The initial learning rate was set to 0.001 and dynamically updated in accordance with the RMSProp formula. Again, we used dropout with a 50% probability and employed an early stopping mechanism that only updates model weights when the loss on the held-out validation set is lower than before.

Both the unidirectional nVAD and the bidirectional decoding model were implemented within the PyTorch framework. For LPCNet, we used the C-implementation and pretrained model weights by the original authors and communicated with the library via wrapper functions through the Cython programming language.

### Closed-Loop Architecture

Our closed-loop architecture was built upon the ezmsg, a general-purpose framework which enables the implementation of streaming systems in the form a directed acyclic network of connected units, which communicate with each other through a publish/subscribe software engineering pattern using asynchronous coroutines. Here, each unit represents a self-contained operation which receives many inputs, and optionally propagates its output to all its subscribers. A unit consists of a settings and state class for enabling initial and updatable configurations and has multiple input and output connection streams to communicate with other nodes in the network. Figure 4 shows a schematic overview of the closed-loop architecture. ECoG signals were received by connecting to BCI2000 via a custom ZeroMQ (ZMQ) networking interface that sent packages of 40 ms over the TCP/IP protocol. From here, each unit interacted with other units through an asynchronous message system that was implemented on top of a shared-memory publish-subscribe multi-processing pattern. Figure 4 shows that the closed-loop architecture was comprised of 5 units for the synthesis pipeline, while employing several additional units that acted as loggers and wrote intermediate data to disc.

In order to play back the synthesized speech during closed-loop sessions, we wrote the bytes of the raw PCM waveform to standard output (stdout) and reinterpreted them by piping them into SoX. We implemented our closed-loop architecture in Python 3.10. To keep the computational complexity manageable for this streamlined application, we implemented several functionalities, such as ringbuffers or specific calculations in the high-gamma feature extraction, in Cython.

### Contamination Analysis

Overt speech production can cause acoustic artifacts in electrophysiological recordings, allowing learning machines such as neural networks to rely on information that is likely to fail once deployed – a phenomenon widely known as Clever Hans (Lapuschkin, 2019). We used the method proposed by Roussel et al. (Roussel, 2020) to assess the risk that our ECoG recordings had been contaminated. This method compares correlations between neural and acoustic spectrograms to determine a contamination index which describes the average correlation of matching frequencies. This contamination index is compared to the distribution of contamination indices resulting from randomly permuting the rows and columns of the contamination matrix – allowing statistical analysis of the risk when assuming that no acoustic contamination is present.

For each recording day among the train, test and validation set, we analyzed acoustic contamination in the high-gamma frequency range. We identified 1 channel (Channel 46) in our recordings that was likely contaminated during 3 recording days (D_5_, D_6_, and D_7_), and we corrected this channel by taking the average of high-gamma power features from neighboring channels (8-neighbour configuration, excluding the bad channel 38). A detailed report can be found in Supplementary Figure 1, where each histogram corresponds to the distribution of permuted contamination matrices, and colored vertical bars indicate the actual contamination index, where green and red indicate the statistical criterion threshold (green: p > 0.05, red: p ≤ 0.05). After excluding the neural data from channel 46, Roussel’s method suggested that the null hypothesis could be rejected, and thus we concluded that no acoustic speech has interfered with neural recording.

### Listening Test

We conducted a forced-choice listening test similar to Herff et al. (Herff, 2019) in which 21 native English speakers evaluated the intelligibility of the synthesized output and the originally spoken words. Listeners were asked to listen to one word at a time and select which word out of the six options most closely resembled it. Here, the listeners had the opportunity to listen to each sample many times before submitting a choice. We implemented the listening test on top of the BeaqleJS framework (Kraft, 2014). All words that were either spoken or synthesized during the 3 closed-loop sessions were included in the listening test, but were randomly sampled from a uniform distribution for unique randomized sequences across listeners. Supplementary Figure 3 provides a screenshot of the interface with which the listeners were working.

All human listeners were only recruited through indirect means such as IRB-approved flyers placed on campus sites and had no direct connection to the PI. Anonymous demographic data was collected at the end of the listening test asking for age and preferred gender. Overall, recruited participants were 23.8% male and 61.9% female (14% other or preferred not to answer) ranging between 18 to 30 years old.

### Statistical Analysis

Original and reconstructed speech spectrograms were compared using Pearson’s correlation coefficients for 80 mel-scaled spectral bins. For this, we transformed original and reconstructed waveforms into the spectral domain using the short-time Fourier transform (window size: 50 ms, frameshift: 10 ms, window function: Hanning), applied 80 triangular filters to focus only on perceptual differences for human listeners (Stevens, 1937), and Gaussianized the distribution of the acoustic space using the natural logarithm. Pearson correlation scores were calculated for each sample by averaging the correlation coefficients across frequency bins. The 95% confidence interval (two-sided) was used in the feature selection procedure while the z-criterion was Bonferroni corrected across time points. Lower and upper bounds for all channels and time points can be found in the supplementary data. Contamination analysis is based on permutation tests that use t-tests as their statistical criterion with a Bonferroni corrected significance level of α = 0.05 / N, where N represents the number of frequency bins multiplied by the number of selected channels.

Overall, we used the SciPy stats package (version 1.10.1) for statistical evaluation, but the contamination analysis has been done in Matlab with the statistics and machine learning toolbox (version 12.4).

### Supplementary Video

This study is accompanied by a video of the participant during one block of a closed-loop session, which demonstrates identification of speech segments from the participant through the unidirectional voice activity detection RNN and closed-loop reconstruction of the spoken speech signal. Note that we masked out the delayed feedback from the patient’s speech, which was recorded from the microphone while played back on the loudspeaker. Instead, we incorporated the same acoustic signal that was played back on the loudspeaker as a separate channel described as BCI (dark red).

## Data Availability

All data will be made available after being accepted in a Journal.

## Acknowledgements

This clinical trial is funded by a BRAIN Initiative UH3 NS114439 from NINDS (PI N.C., co-PI N.R).

## Author Contributions

M.A. and N.C. wrote the manuscript. M.A., S.L., Q.R. and D.C. analyzed the data. M.A. and S.S. conducted the listening test. S.L. collected the data. M.A. and G.M. implemented the code for the online decoder and the underlying framework. M.A. made the visualizations. W.A., C.G. and K.R., L.C. and N.M. conducted the surgery / medical procedure. F.T. handled the regulatory aspects. H.H. supervised the speech processing methodology. M.F. N.R. and N.C. supervised the study and the conceptualization. All authors reviewed and revised the manuscript.

## Competing Interests

The authors declare that they have no competing interests.

## Data and Code Availability

Neural data and anonymized speech audio will be made publicly available on www.osf.io upon acceptance of the manuscript. Corresponding source code for the closed-loop BCI and scripts for generating figures can be obtained from the official Crone Lab Github page at: https://github.com/cronelab/… and will be made publicly available upon acceptance of the manuscript. The ezmsg framework can be obtained from https://github.com/iscoe/ezmsg.

## Supplementary Materials

**Supplementary Figure 1.**
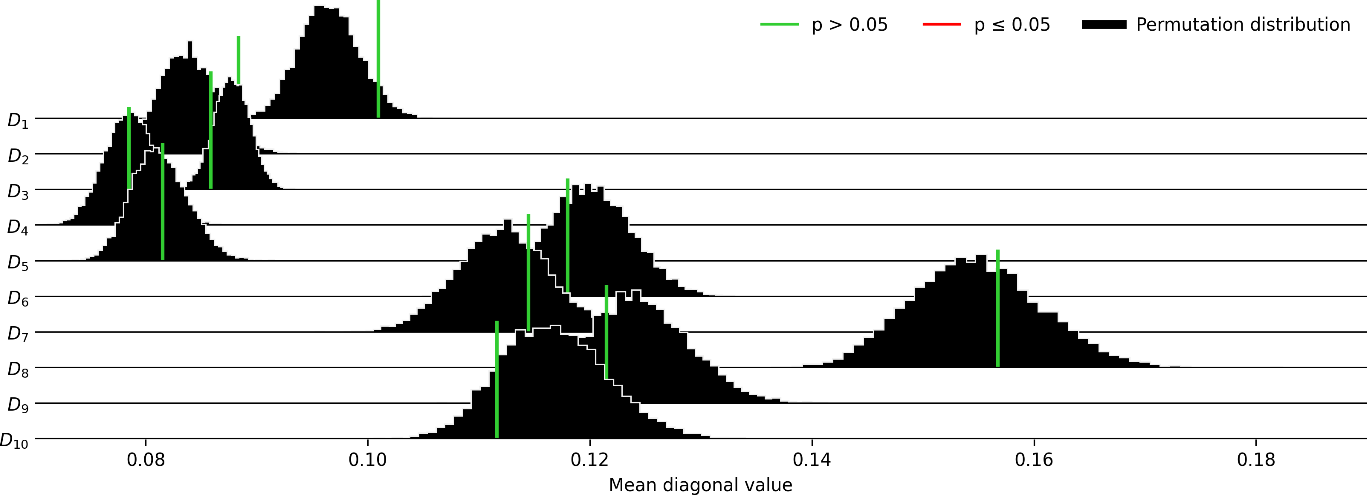
| Acoustic contamination report. We checked all recordings in our training, validation and test sets for acoustic contamination using the method proposed by Roussel et al. (Roussel, 2020). For each day, the histogram represents mean diagonal values of the distribution of permutated contamination matrices (N=10,000). Colored bars indicate the position of the contamination index of neural recordings, while the statistical criterion for rejecting the null hypothesis is indicated in green (p > 0.05) or in red (p ≤ 0.05) if acoustic contamination is present. After correcting Channel 46 for D_5_, D_6_, and D_7_ we did not detect any acoustic contamination. Recordings of the closed-loop sessions where not checked for acoustic contamination since they were completely withheld from network training procedures, and therefore cannot leak into model training and lead to the Clever Hans phenomenon (Lapuschkin, 2019).

**Supplementary Figure 2.**
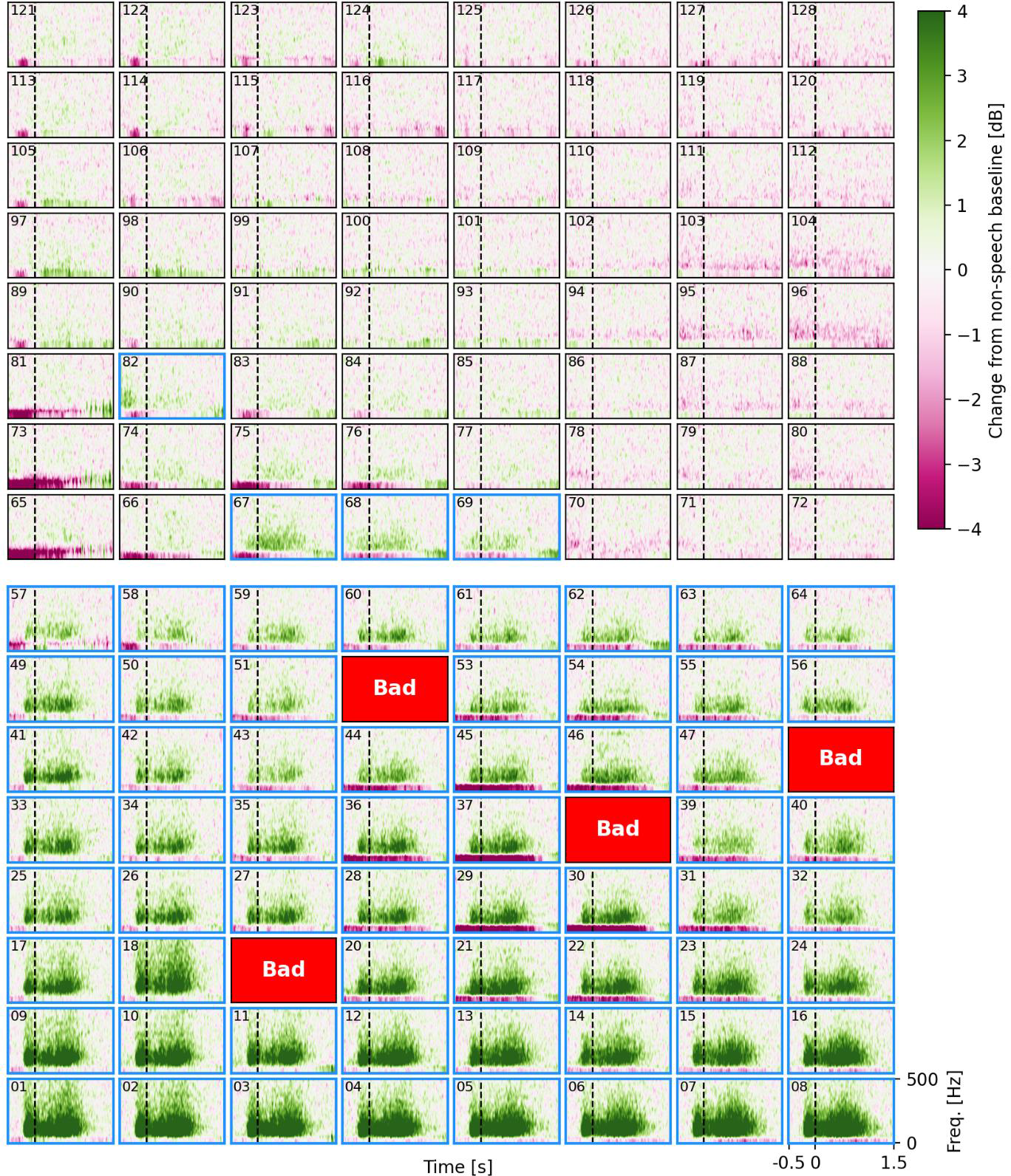
| Power spectral analysis for identifying speech-related channels. Each plot shows channel specific normalized power spectral density in the frequency range of 0 - 500 Hz averaged across all keywords for a particular recording session in the training set (60 trials). Dashed lines indicate speech onset times, while the x-axis shows time-aligned power spectral density in the range of −0.5 s up to 1.5 s. Bad channels were excluded based on visual inspection. Channels marked with blue borders represent the top-64 channels whose lower bound of a 95% confidence interval most often exceed baseline high-gamma activity (see Supplementary Data for upper and lower bounds, training set only) and were selected as relevant channels for the speech synthesis task.

**Supplementary Figure 3.**
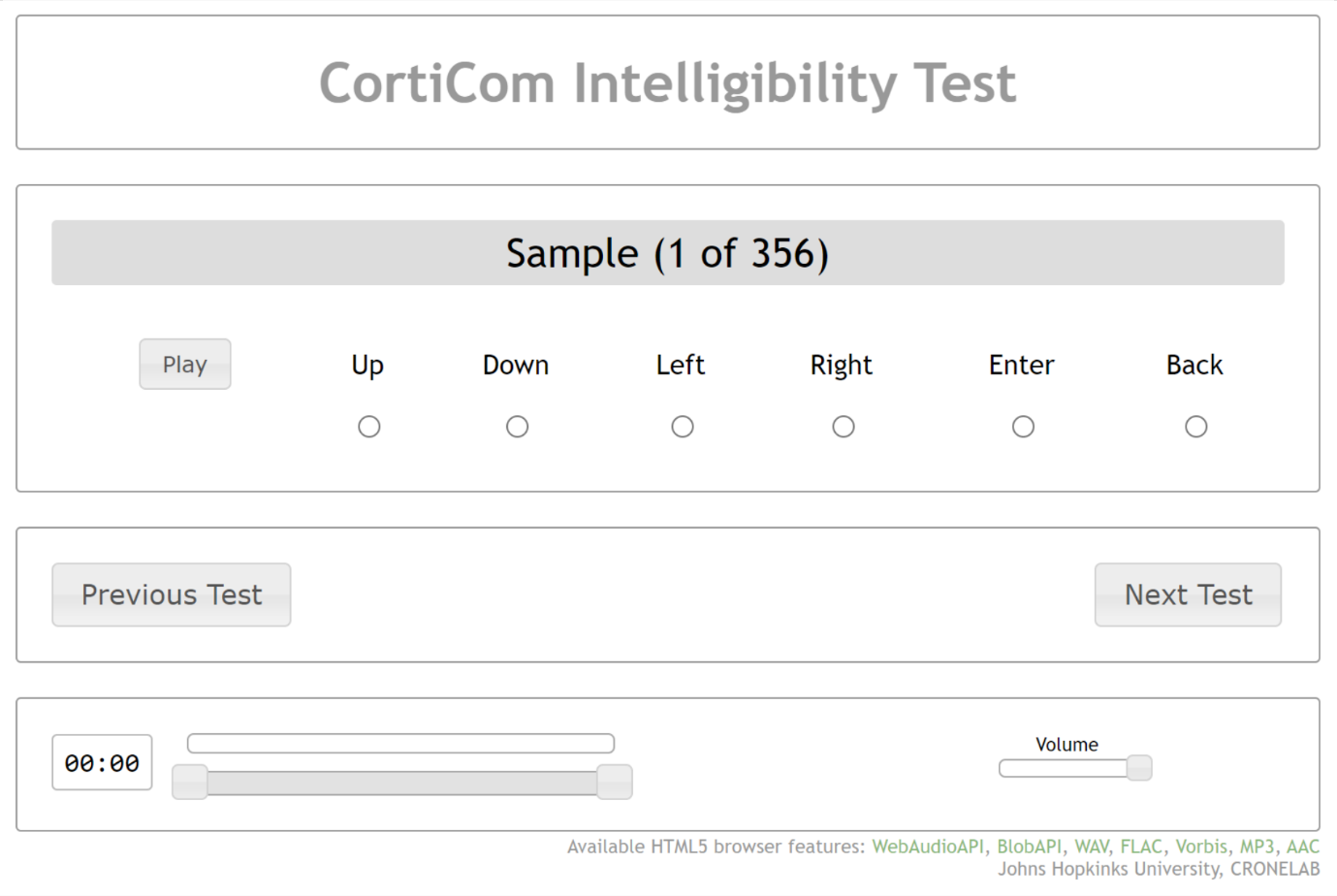
| Web-Interface for conducting the listening test. All synthesized samples are presented in a randomized sequence. Human listeners can use the Play button to play the sample and subsequently select one of the radio buttons to make their choice or play the sample again. After making a choice they can move on to the next sample. Human listeners can go back to the previous sample if they accidentally make a wrong decision, but not beyond it.

**Supplementary Table 1.**
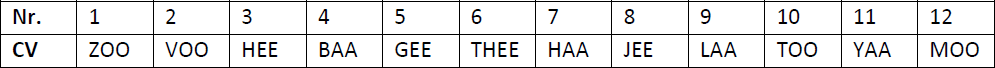
| Syllable stimuli from the syllable repetition task. Based on Bouchard et al. (Bouchard, 2013) we chose 12 consonant-vowel (CV) syllable stimuli to acquire high-gamma activity of articulatory representations for a stable baseline across days.

## Notes

### Competing Interest Statement

The authors have declared no competing interest.

### Clinical Trial

NCT03567213

### Funding Statement

This study was funded by a BRAIN Initiative UH3 NS114439 from NINDS

### Author Declarations

The Institutional Review Board (IRB) of the Johns Hopkins University gave ethical approval for this work and the Food and Drug Administration (FDA) gave approval under an investigational device exemption (IDE)

